# Pregnancy-induced changes in blood composition drive post-partum hemorrhage risk

**DOI:** 10.1101/2021.05.14.21257156

**Authors:** Matthew R. Robinson, Marion Patxot, Miloš Stojanov, Sabine Blum, David Baud

## Abstract

The extent to which women differ in the course of blood cell counts throughout pregnancy, and the importance of these changes to pregnancy outcomes has not been well defined. Here, we develop a series of statistical analyses of repeated measures data to reveal the degree to which women differ in the course of pregnancy, predict the changes that occur, and determine the importance of these changes for post-partum hemorrhage which is one of the leading causes of maternal mortality. We present a prospective cohort of 4,082 births recorded at the University Hospital, Lausanne, Switzerland between 2009 and 2014 where full labour records could be obtained, along with complete blood count data taken at hospital admission. We find significant differences, at a *p* < 0.001 level, among women in how blood count values change through pregnancy for mean corpuscular hemoglobin, mean corpuscular volume, mean platelet volume, platelet count and red cell distribution width. We find evidence that almost all complete blood count values show trimester-specific associations with postpartum hemorrhage and that tracking blood count value changes through pregnancy improves identification of women at increased risk, with increased area under the receiver operator curve in independent patient samples. Differences among women in the course of blood cell counts throughout pregnancy have an important role in shaping pregnancy outcome. Modelling trimester-specific associations with pregnancy outcomes, in a way that fully utilizes repeated measures data, provides greater understanding of the complex changes in blood count values that occur through pregnancy and provides indicators to guide the stratification of patients into risk groups.

## Introduction

Complete blood cell count (CBC) values measured during the course of pregnancy can provide an indicator of a range of clinical risk factors and birth outcomes^1–4^. A number of studies describe the typical ranges of CBC measurements observed during the course of pregnancy, characterising the differences observed among women, and allowing identification of women with abnormal measurements for follow-up^1,2,5–7^. However, women differ in how their physiology changes throughout pregnancy, and observing a large change in CBC value relative to a previous measurement (a within-individual comparison), may be more indicative of pregnancy outcome, than only a single observation compared to a baseline range (a between-individual comparison). In this work, we propose that characterising the changes that occur within women over the course of their pregnancy and relating these changes to pregnancy outcomes, may improve understanding of the course of pregnancy and facilitate better patient screening.

Here, we demonstrate that changes in CBC value relative to previous measurements are more relevant to the prediction of post-partum hemorrhage (PPH) than measures taken at a single point in time. PPH is one of the leading causes of maternal death worldwide^8,9^. Despite its ubiquity, the root causes of PPH remain obscure^10^. In a typical delivery, uterine bleeding at the site of placental separation from the uterine wall is stopped by constriction of the blood vessels due to the uterine muscle contraction and by clots sealing off the placental bed. When uterine bleeding is not stopped by these mechanisms, the threat of maternal death from catastrophic hemorrhage is high with clinical PPH defined as a loss of approximately 500 cubic centimeters of blood at or around delivery (for vaginal births). The underlying causes of uterine atony, however, are unclear, with a number of risk factors proposed that so far, fail to provide early pregnancy indicative measures. In a prospective cohort of 4,082 births, we show how improved statistical analysis can reveal the degree to which women differ in CBC value through the course of pregnancy, predict the changes that occur, determine the importance of these changes for a clinical outcome that is one of the leading causes of maternal mortality, and potentially provide improved early-pregnancy screening for labour outcomes.

## Results

### INDIVIDUAL TRACKING OF BLOOD COUNT VALUE CHANGE

Data on self-reported ethnicity was collected, with women describing their country of origin. The sample is ethnically diverse, with 68% of individuals reporting their origin as being a European country, with the remaining 32% reporting one of 62 different non-European origin countries. The mean age of the women at birth was 30.65, the minimum was 18 and the maximum was 48. Through all analyses presented below, the country of origin of the women was fitted as an additional covariate to control for ancestry differences.

For 1,457 women with complete blood count measures recorded in each of their three trimesters, we find differences among women in how blood count values change through pregnancy for six CBC measures: red cell distribution width (RDW), mean corpuscular hemoglobin (MCH) and it’s concentration (MCHC), mean corpuscular volume (MCV), platelet count (PLATE), and mean platelet volume (MPV; Figure 1). We modelled the data using a repeated measures model to estimate an intercept parameter, describing the woman’s ‘initial’ blood count value at the start of pregnancy. The proportion of variance attributable to this term gives the individual repeatability, the extent to which observations on the same individuals are correlated across trimesters. We also estimated the slope, or rate of change parameter, describing the woman’s rate of change over time, with the proportion of phenotypic variance attributable to the rate of change term, describing the magnitude of the differences among women in the course of blood cell counts throughout pregnancy.

**Figure 1.**
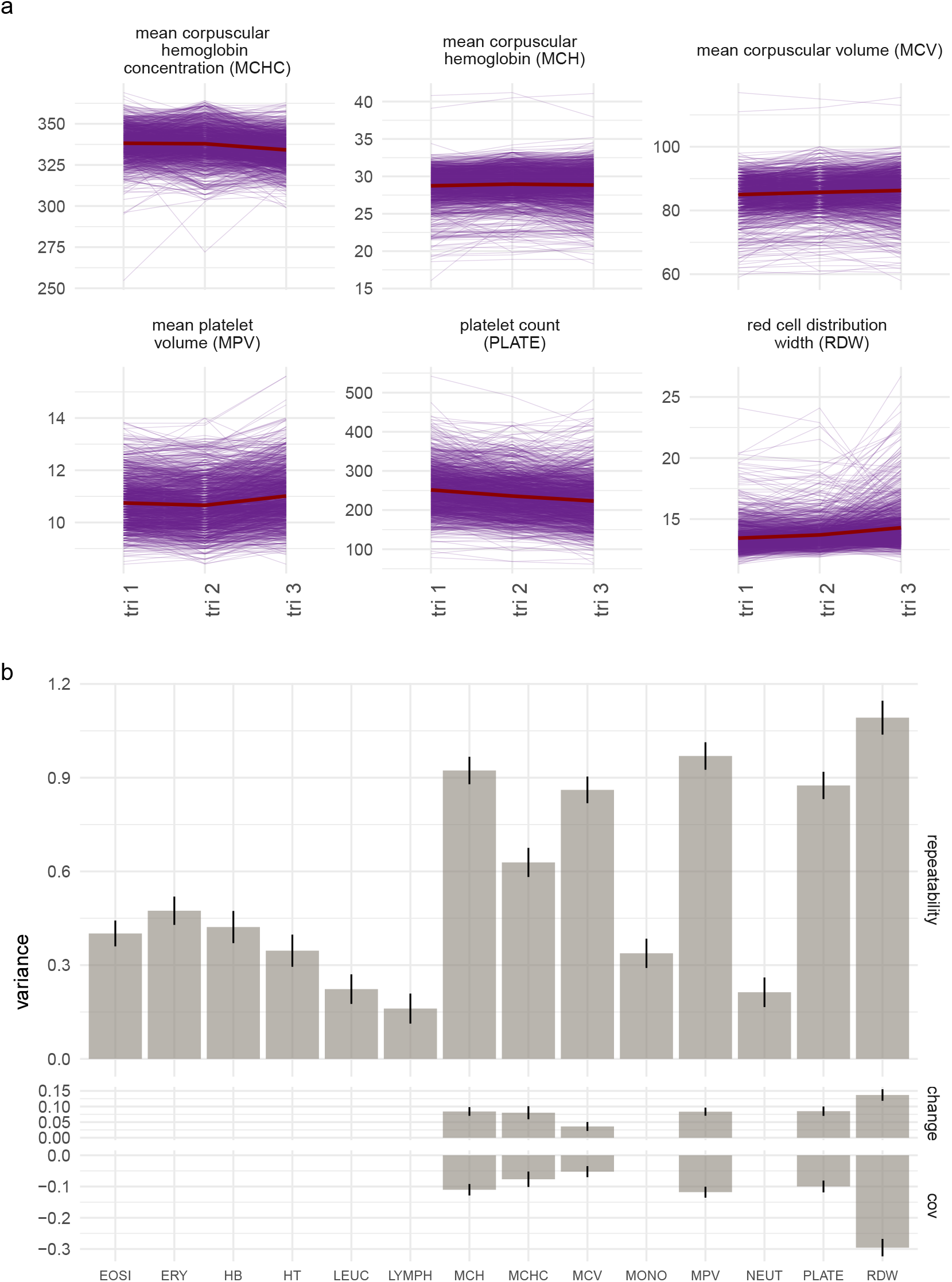
Tracking the complete blood cell count values of women across pregnancy. (a) We find seven blood cell count measures for which there are significant differences among women in how their values change through pregnancy and we present the main six here. Lines represent the values recorded across three trimesters for each individual, with the red line giving the population mean. (b) We modelled the data using a repeated measures model to estimate individual repeatability, defined as the proportion of observed variance attributable to among-individual differences, and then to determine whether women differ in the changes that occur in blood cell counts across pregnancy, by estimating the proportion of observed variance attributable to among-individual differences in the changes of the blood count values across the three trimesters (labelled “change”). The parameter estimates from the repeated measures model are shown with error bars giving the SE, with the covariance between the individual-level differences and their changes labelled cov. The repeatability gives the extent to which observations on the same individuals are correlated across trimesters. The change gives the extent to which women differ in the change or trajectory of measures across pregnancy. If there was no evidence at a *p* < 0.001 level for a model that includes among-women changes, then only the repeatability model estimate is given. Blood count measure abbreviations: erythrocytes (ERY, red blood cells), hematocrit levels (volume percentage of red blood cells, HT), mean corpuscular hemoglobin (MCH) and mean corpuscular hemoglobin concentration (MCHC), mean corpuscular volume (MCV), hemoglobin levels (HB), red cell distribution width (RDW), platelets (PLATE), mean platelet volume (MPV), leucocytes (LEUC), eosinophils (EOSI), lymphocytes (LYMPH), monocytes (MONO), and neutrophils (NEUT)

We find variation among the blood count measures in how women differ in the changes that occur across pregnancy (Figure 1, Figure **??**). Erythrocye count (ERY), hemoglobin levels (HB), leucocyte (LEUC) count, absolute counts (the percentage of cells multiplied by the LEUC count) of lymphocytes (LYMPH), monocytes (MONO), and neutrophils (NEUT) show a correlation across trimesters of less than 0.5, with no evidence of consistent differences among women in how the values change across pregnancy (Figure 1, Figure **??**). Thus for these measures, values fluctuate considerably and early pregnancy measures do not strongly relate to (are not predictive of) measurements repeated later in pregnancy.

In contrast, absolute eosinophil count (EOSI), show high correlation across measures and no evidence for differences among women in the changes of these values across pregnancy (Figure 1, Figure **??**). This implies that EOSI measures are consistent, with early pregnancy measures being informative as to later pregnancy values, with all women responding the same way across pregnancy.

For platelet count (PLATE), mean corpuscular volume (MCV), mean platelet volume (MPV), mean corpuscular hemoglobin mass (MCH) and concentration (MCHC), and red cell distribution width (RDW) we find high repeatability and evidence for differences among women in the changes that occur as pregnancy proceeds (Figure 1, Figure **??**). Thus for some women their values will increase, whereas for others their values will decrease through pregnancy, with early measures being indicative of both the later values and the changes that occur. These results show that fully utilizing repeated measures through pregnancy by directly modelling individual-level change, provides a characterisation of the course of blood cell counts throughout pregnancy for each woman, that can be used to predict the trajectories of women through their pregnancy.

### ASSOCIATIONS OF BLOOD COUNT VALUE CHANGES AND PPH

We find evidence that complete blood count values show trimester-specific associations with PPH (Figure 2). We use a Bayesian path coefficient model that jointly estimates relationships among repeated blood count measures across trimesters, as well as the association of each trimester to a binary (0/1) outcome variable adjusting for the values of the others trimesters. The outcome - blood measure associations obtained are thus specific to the changes that occur during pregnancy, with trimester 1 associations reflecting the baseline association of the values women present at the start of their pregnancy; trimester 2 associations reflecting the association of trimester 2 values adjusting for the values of each women at trimester 1; and trimester 3 associations reflecting the association of trimester 3 values adjusting for the values of each women at trimester 1 and 2.

**Figure 2.**
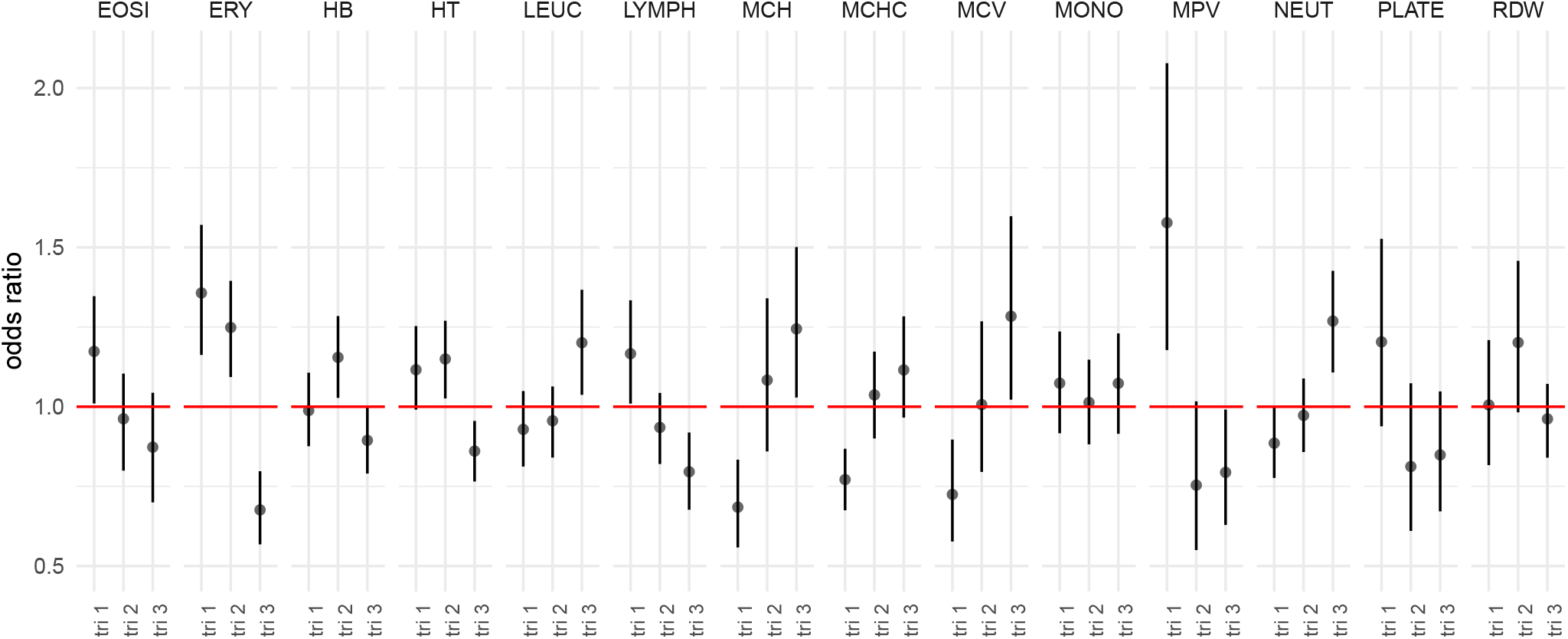
Trimester-specific associations of CBC values and PPH outcome. Odds ratios for complete blood count measures (CBC) recorded at each of the three trimesters (tri 1, tri 2, tri 3) for an indicator variable of whether the women lost ≥ 500ml of blood post-partum (PPH). Values are the posterior mean, with error bars showing the 95% credible intervals, obtained from a Bayesian path coefficient model that jointly estimates relationships among blood count measures and their relationships to a binary (0/1) outcome variable. Results are shown for blood counts where the 95% credible intervals were different to odds ratio 1 for at least one trimester measure. Blood count measure abbreviations: erythrocytes (ERY, red blood cells), hematocrit levels (volume percentage of red blood cells, HT), mean corpuscular hemoglobin (MCH) and mean corpuscular hemoglobin concentration (MCHC), mean corpuscular volume (MCV), red cell distribution width (RDW), platelets (PLATE), mean platelet volume (MPV), leucocytes (LEUC), eosinophils (EOSI), lymphocytes (LYMPH), monocytes (MONO), and neutrophils (NEUT)

Women whose average leukocyte count was high in the last trimester are at increased risk of postpartum hemorrhage (Figure 2). For the white blood cells that compose the leukocyte count, we find women with increasing neutrophil white blood cell composition in trimester three relative to earlier pregnancy is associated with increased risk of postpartum hemorrhage (Figure 2). This suggests that elevated phagocytic cell composition in early pregnancy is likely indicative of an underlying immune disease, with increasing levels in the third trimester indicative of the occurrence of infection (mainly chorioamnionitis), known to decrease coagulation.

High eosinophil and lymphocyte composition within the white blood cell count at baseline (trimester 1) is associated with increased risk of PPH (Figure 2). This suggests that an early-pregnancy inflation-associated immune response may hinder placental development, resulting in higher later-pregnancy risk of PPH. However, women whose white blood cell count compositions change with increased eosinophil and lymphocyte composition in the third trimester have reduced risk of postpartum hemorrhage (Figure 2). Changes in blood vessel permeability so that neutrophils and clotting proteins can get into connective tissue more easily may reflect the normal pregnancy progression in immune inflammation response, with women who present greater changes than the average being those least at risk of PPH.

We find that associations of postpartum hemorrhage and erythocyte count and total hematocrit and heamoglobin levels to vary across pregnancy, with women whose values increased in the first and second trimester, showing increased risk of PPH (Figure 2). This provides better understanding of previous epidemiologic studies that have also found an association between high maternal hemoglobin concentrations and an increased risk of poor pregnancy outcomes. This association may be attributable to hypertensive disorders of pregnancy and to preeclampsia as the pathophysiologic mechanism of these conditions can produce higher hemoglobin concentrations through reduced plasma expansion and reduced placental-fetal perfusion.

We also find that high mean corpuscular hemoglobin, and mean corpuscular volume in later pregnancy relative to earlier measures are associated with increased likelihood of PPH, which likely indicates folate and B12 deficiency (Figure 2).

Generally, high platelet count and high mean platelet volume later are associated with reduced the risk of PPH (Figure 2). High platelet volume at trimester 1 is associated with increased risk of PPH, however increasing levels throughout pregnancy relative to this first measure decrease PPH risk (Figure 2). This supports previous results that platelet counts may be lower in women with pregnancy-related complications, and it also lends support to previous studies showing that for early-pregnancy hypertension a significant higher platelet volume may indicate the severity of disease. This highlights that early diagnosis of progressive activation of coagulation may help manage PPH, with platelet activation markers a useful guide alongside other CBC values.

### IMPROVED OUTCOME PREDICTION THROUGH BLOOD COUNT VALUE TRACKING

The associations of postpartum hemorrhage with changes in complete blood cell count values represent substantial odds ratios (Figure 2), implying that tracking complete blood cell count value through pregnancy, can improve understanding of the incidence of later pregnancy outcomes and improve prediction of outcome. We set out to test this by first modelling relationships between PPH and complete blood cell counts taken at the three trimesters as compared to tracking women through their pregnancy. Within repeated held-out set of 200 patients, the average AUC increases from the baseline when using CBC measures from trimester three, and increases further when using CBC values recorded at three time points across pregnancy (Figure 3). This demonstrates that tracking blood count value changes through pregnancy improves identification of women who are at increased risk of PPH.

**Figure 3.**
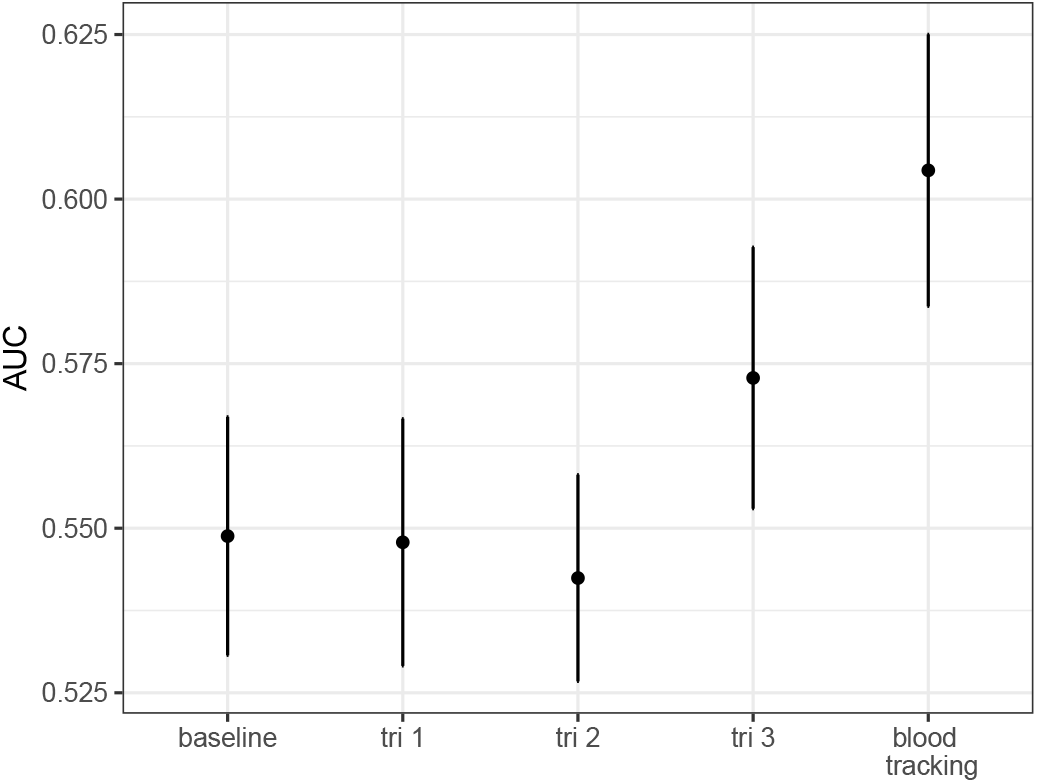
Comparison of model predictive performance. Mean and SE of the area under the receiver operator curve (AUC) for repeatedly held out samples of 200 independent patients from a model including only age and ancestry (baseline), baseline and complete blood count (CBC) measures taken at trimester 1 (tri 1), baseline and complete blood count (CBC) measures taken at trimester 2 (tri 2), baseline and complete blood count (CBC) measures taken at trimester 3 (tri 3), and baseline and complete blood count (CBC) measures taken across pregnancy (blood tracking).

## Discussion

Our analysis provides evidence that not only platelet counts, but also blood hemoglobin levels, hemoglobin concentrations and immune response plays a vital role in shaping the underlying risk of PPH. Associations of postpartum hemorrhage with changes in complete blood cell count values represent substantial odds ratios, and investigating these processes will provide improved understanding of the incidence of later pregnancy outcomes and improve prediction of outcome.

In humans, extensive vascular remodeling must occur to allow for placentation and establishment of early pregnancy, as well as to support the demands of a growing fetus. Not only is adequate vascular remodeling critical for the establishment of a normal pregnancy, but abnormalities in these early events are associated with later complications of pregnancy such as preeclampsia and intrauterine growth restriction, which can have a major impact on fetal and neonatal health. In general, pregnancy hormones are thought to suppress maternal alloresponses, while promoting pathways of tolerance. Each of the major pregnancy-associated hormones is thought to directly and indirectly affect the function of the major immune cells and thus impacts the immune milieu during pregnancy. The general pregnancy response has been described as an increased neutrophil and monocyte count, whereas lymphocytes, and eosinophils declined in number. Here, we show that differences in women in the rates of these changes are important, with lower declines in lymphocytes and eosinophils throughout pregnancy and lower increases in neutrophil and monocyte count associated with reduced risk of postpartum hemorrhage. Thus, maintaining ‘steady-state’ immune cell levels may be key to a healthy pregnancy.

Although gestational thrombocytopenia has been recognized for more than 25 years, the degree of differences among women in the course of platelet counts throughout pregnancy has not been described, nor have the implications been assessed. Dilution of platelets may occur simply due to the increased plasma volume that occurs during pregnancy. Too limited plasma dilution early in pregnancy giving a higher platelet count, may be indicative of other early-pregnancy disorders, which then increase the risk of postpartum hemorrhage. Evidence suggests that placental circulation is similar to splenic circulation, and thus platelets may accumulate within the intervillous space of the placenta and this accumulation may be key to limiting postpartum hemorrhage later in pregnancy. Thus, platelet activation markers are a useful guide for the early diagnosis of progressive activation of coagulation, which may help manage postpartum hemorrhage risk in conjunction with other blood measures.

We have shown how modelling trimester-specific associations with pregnancy outcomes in a way that fully utilizes repeated measures data, can provide greater understanding of the complex changes in blood count values that occur through pregnancy and provide indicators to guide the stratification of patients into risk groups early in their pregnancy, potentially facilitating improved care. These association represent hypothesis that now require further work to understand. Combining other sources of repeated measures with other risk factor information is also likely to be important for creating improved patient prediction, and this is a focus of future work. We provide an algorithm enabling practitioners to record repeated CBC measures and to predict PPH prior to labour at: https://medical-genomics-group.shinyapps.io/PredPPH/

Other factors such as induction of labor, prolonged labor, instrumental delivery and chorioamnionitis, are important for the risk of PPH, but these are only known shortly before or during birth and the focus of this study is not the generation of the highest AUC, but rather a demonstration that repeated measures data through pregnancy can be utilized to predict patient outcomes in advance of delivery. Our predictor remains limited in its application, and this is solely a function of the sample size of the study presented here. Large-scale data collection to create a comprehensive, repeatedly collected medical record, will improve prediction ability for future patients.

In summary, our repeated measures analyses demonstrates that women differ in the course of blood cell counts throughout pregnancy, and that these differences have an important role in shaping pregnancy outcomes.

## Methods

### STUDY POPULATION

This study is composed of 5,205 births recorded at the University Hospital, Lausanne, Switzerland between 2009 and 2014, for which ethics consent was obtained from the local Commission cantonale (VD) d’ethique de la recherche sur l’être humain (CER-VD), project-ID 2019-00280). All data were collected prospectively at the time of delivery by the obstetrician or midwife. We previously cross-checked our database, with confirmation of congruent data in >98% of cases^11^. For 4,082 of the births, full labour records could be obtained, along with complete blood count data taken at hospital admission. All research was performed in accordance with relevant guidelines/regulations. Informed consent was obtained from mothers for use of their data and from mothers for use of the data on their children.

### COMPLETE BLOOD COUNT VALUES

All 4,082 births had complete blood count data recorded at hospital admission prior to labour. The blood cell count data consisted of absolute counts of erythrocytes (ERY, red blood cells), hematocrit levels (volume percentage of red blood cells, HT), mean corpuscular hemoglobin (MCH) and mean corpuscular hemoglobin concentration (MCHC), mean corpuscular volume (MCV), hemoglobin levels (HB), red cell distribution width (RDW), platelets (PLATE), mean platelet volume (MPV), leucocytes (LEUC), eosinophils (EOSI), lymphocytes (LYMPH), monocytes (MONO), and neutrophils (NEUT).

For 1,457 of these births, women had complete blood count data recorded during all three trimesters (0 to 13 weeks, 14 to 27 weeks, and 28 to 42 weeks). To determine the course of complete blood count values throughout pregnancy, we took the average of values collected within each trimester for each woman, so that each woman had three repeated measures.

### STATISTICAL ANALYSIS

#### Multiple imputation of the data

Under the assumption that data is missing at random, within the data set of 4,082 births, and within the data set of 1,457 women we imputed missing data using the R package mice. We used standard procedures and ensured no variable had greater than 15% missing rate. We generated three repeated sets of complete data, analysing each one and then averaging the results.

#### Modelling the course of blood cell counts throughout pregnancy for each woman

We first modelled the repeated measures across the three trimesters using a random coefficient model (also known as a latent growth model), implemented in the R package lavaan. The individual change trajectory can be described by a ‘within-person’ or ‘level-1’ individual growth model. We hypothesise that the blood count value of measure *k*, recorded at trimester *j*, for women *i* is a linear function of time *t*

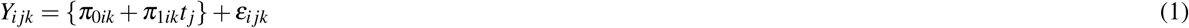

where *i* = 1, 2,, *N*, with *N* the number of individuals, *j* = 1, 2, 3, and *t*_1_ = 0, *t*_2_ = 1, *t*_3_ = 2. *ε*_*i jk*_ are the measurement occasion specific errors which are assumed to be independent for the individual growth parameters *π*_0*ik*_ and *π*_1*ik*_. *π*_0*ik*_ is the intercept parameter, describing the woman’s ‘initial’ blood count value of measure *k* at the start of pregnancy as it is defined as the origin of time. *π*_1*ik*_ is the slope, or rate of change parameter, describing the woman’s rate of change over time. Here, while each woman’s change is modelled with the same functional form (linear), women have different values of the slope parameters. Thus, if there is variance in *π*_1*ik*_, than women differ women differ in the course of blood cell counts throughout pregnancy. A linear form represents a modelling choice and we did not extend the model further to test for non-linear variation. This is because we simply wished to establish whether there are individual-level differences in changes across pregnancy and we did not feel that the present sample size of this study was sufficient for testing specific non-linear forms of change.

In matrix notation for each blood count measure, dropping subscript *k*:

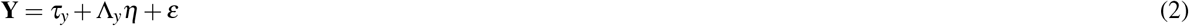

where **Y** is a vector of values of the *k*^*th*^ observed blood count, *τ*_*y*_ is a vector containing the population means of **Y**, Λ_*y*_ is a matrix of factor loadings, *η* is a vector of latent variables, and *ε* is a vector of residuals. So with three repeated measures the matrices are

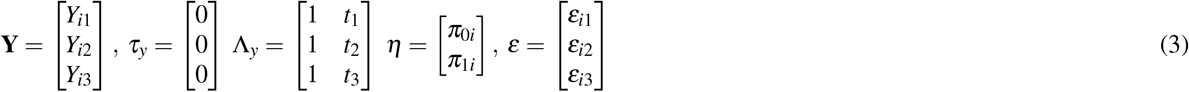

We ran this model for each of the 14 complete blood cell count measures and estimated the proportion of phenotypic variance attributable to *π*_0*ik*_ and *π*_1*ik*_.

#### Association of blood cell counts and PPH through pregnancy

We then wished to assess the unique contribution of differences among women in their complete blood counts values at each trimester to their risk of postpartum hemorrhage.

We restricted the data to those women where CBC values could be linked to information on the occurrence of PPH. For each delivery in our institution, graduated collector bags were placed just after birth, left in place at least for 15 minutes, and then until the birth attendant judged that bleeding had stopped. Amount of blood loss was recorded in the medical file. Our management of PPH is described elsewhere and follow international guidelines. We selected women who had delivered a live child through vaginal birth (Cesarean sections were excluded), giving 1,012 births where women had three repeated blood measures across trimesters. PPH was defined as blood loss of 500 ml or more after vaginal birth (n=119 of the 1,012 women).

We used a model known as a path analysis, involving a series of jointly estimated equations

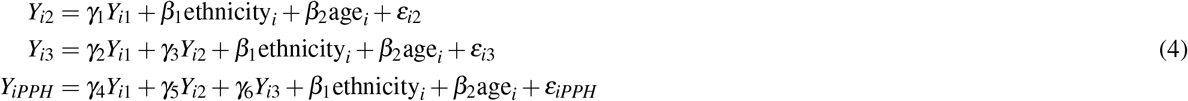

where, *Y*_*i*1_, *Y*_*i*2_ and *Y*_*i*3_ are the complete blood count values for a given measure recorded at the first, second, and third trimester respectively, *Y*_*iPPH*_ is an indicator 0/1 of PPH. This can be represented as the matrix equation

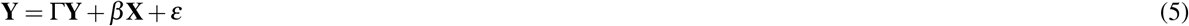

where,

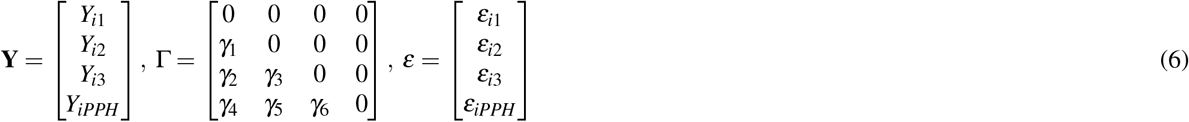

with **X** a design matrix containing the covariates ethnicity and age, and *β* the regression coefficients of these covariates for each measurement. This model estimates relationships among repeated measurements, and determines the unique contribution of each trimester measure on the outcome indicator of postpartum hemorrhage adjusting for the values of previous blood measures. So *γ*_4_ estimates the association of the first trimester blood count value measurements and the outcome, *γ*_5_ estimates the association of the second trimester blood count value measurements and the outcome adjusting for the associations at the first trimester, and *γ*_6_ estimates the association of the third trimester blood count value measurements and the outcome adjusting for all previous trimester associations.

We implemented this model in the R package brms providing Bayesian inference of the model parameters. We modelled *Y*_*PPH*_ as a 0/1 variable using a Bernoulli logit link function, specifying non-informative priors over the model parameters. We used a burnin period of 1000 iterations, sampling for a total of 2500 iterations, across 3 independent chains. We performed full diagnostic and posterior predictive checks to ensure parameter convergence across chains. We determined associations on the basis of whether the 95% credible intervals of the estimates overlapped zero.

#### Patient prediction of PPH

We used a logistic regression implemented in brms, with a Bernoulli logit link function, to model the relationship between the indicator 0/1 of PPH the complete blood count values recorded at hospital admission for labour for the 1,012 births where women had three repeated blood measures across trimesters. For this analysis, we randomly selected a subset of 200 women (175 non-postpartum hemorrhage and 25 postpartum hemorrhage) with complete birth records and removed these from the data. We ran four models in the remaining data:

- a model of the relationship between the indicator 0/1 of PPH and CBC data of the first trimester
- a model of the relationship between the indicator 0/1 of PPH and CBC data of the second trimester
- a model of the relationship between the indicator 0/1 of PPH and CBC data of the third trimester
- a model of the relationship between the indicator 0/1 of PPH and CBC data recorded across the three trimesters

For each model we used a normal prior for the intercept and the regression coefficients, with mean of zero and variance of two, with a burn-in period of 1000 iterations, sampling of 2500 iterations, and three independent chains. For each of the four models, we predicted postpartum hemorrhage outcome in the left-out subset of 200 women and calculated the area under the receiver operator curve. We repeated the sub-sampling, the analysis of the three models, and the prediction ten times and we plot the comparison of the mean area under the receiver operator curve (AUC) for the four models across the ten replicates.

## Data availability

Data are available through joint research agreements from the corresponding authors.

## Data Availability

Data are available through joint research agreements from the corresponding authors.

## Acknowledgements

This project was funded by an SNSF Eccellenza Grant to MRR (PCEGP3-181181), and by core funding from the Institute of Science and Technology Austria. We would like to thank the participants of the study and all the midwives and doctors for the computerized obstetrical data.

## Author contributions statement

MRR and DB conceived and designed the study. MRR conducted the experiments and analysis. MP and MS contributed data to the analysis. MRR and DB wrote the paper. All authors approved the final manuscript prior to submission.

