## Supplemental Figure 1 for "Pregnancy-induced changes in blood composition drive post-partum hemorrhage risk"

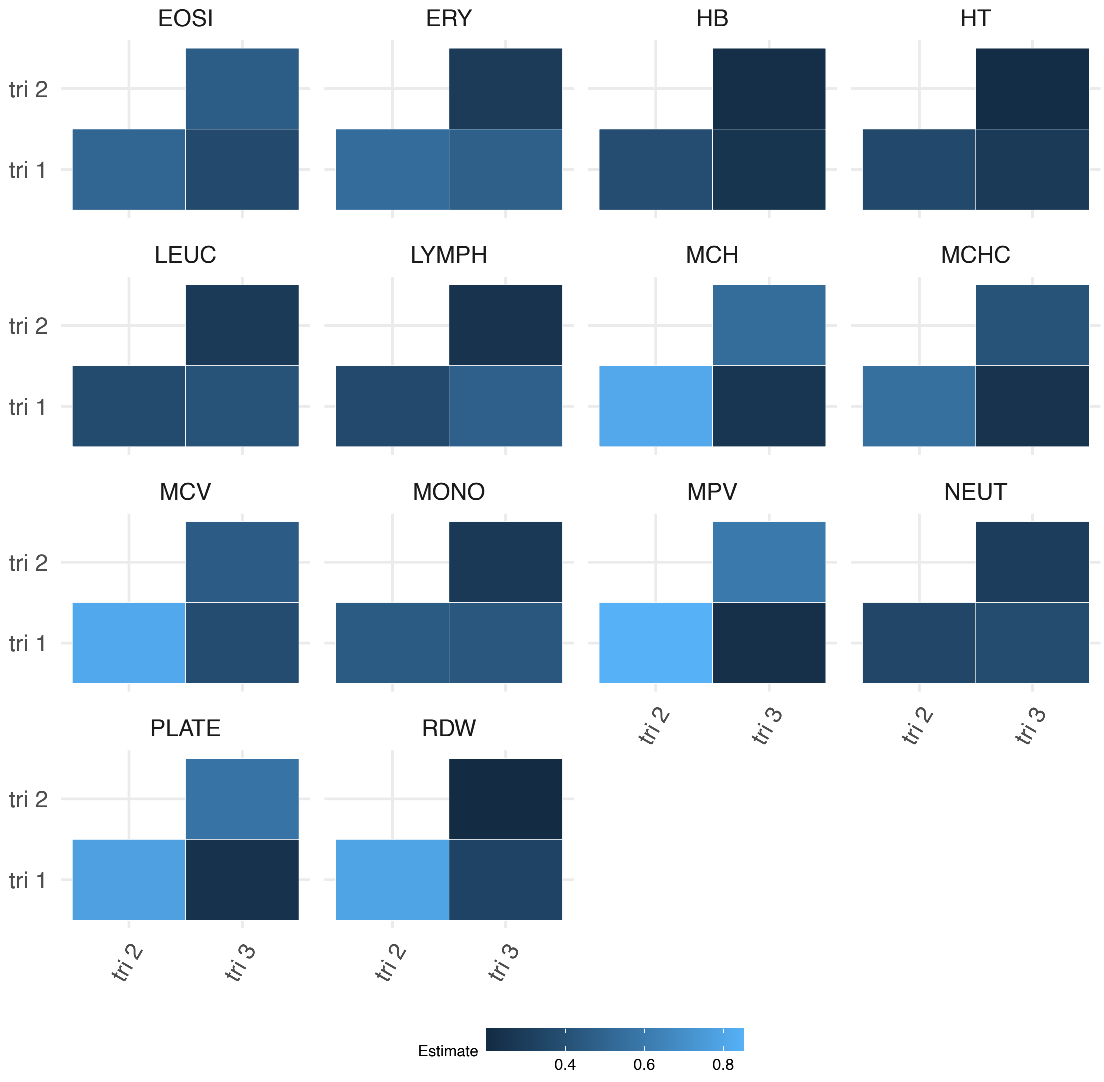


**Figure S1. Path coefficients of trimester-specific complete blood count values**. Path relationships between complete blood count measures indicating the strength of relationship (on the correlation scale) of measures recorded across the three trimesters of pregnancy. Measure abbreviations: absolute eosinophil (EOSI), lymphocyte (LYMPH), monophil (MONO) and neutrolphil (NEUT) levels; leukocyte count (LEUC), erythrocyte count (ERY), hematocrit (HT), hemoglobin level (HB), mean corpuscular hemoglobin (MCH) and the concentration (MCHC), mean corpuscular volume (MCV), mean platelet volume (MPV), platelet count (PLATE), and red cell distribution width (RDW)
